# Development and Validation of Machine Learning Models for Predicting Mortality in Hospitalised Systemic Lupus Erythematosus Patients in Dr. Sardjito Hospital, Indonesia

**DOI:** 10.64898/2026.05.01.26352268

**Authors:** Ayu Paramaiswari, Dhite Bayu Nugroho

## Abstract

**Objectives:** This study aimed to develop and validate machine learning models to predict in-hospital mortality among systemic lupus erythematosus (SLE) patients using administrative claims data in a tertiary referral center in Indonesia.

**Methods:** We conducted a retrospective cohort study of 327 SLE hospital admissions between January 2019 and June 2025. Predictor variables included demographics, hospitalisation characteristics, and the ten most frequent comorbidities. We developed Logistic Regression, Random Forest, and Extreme Gradient Boosting (XGBoost) models. Class imbalance was addressed using the Synthetic Minority Over-sampling Technique.

**Results:** The overall in-hospital mortality rate was 7.7%. While models achieved comparable discrimination (Area Under the Curve ~0.71), XGBoost was selected for its superior sensitivity (0.93) compared to Logistic Regression (0.80) and Random Forest (0.97). Feature importance analysis revealed pneumonia as the most significant predictor, followed by acute kidney failure and length of stay. Hypoalbuminemia and hyponatremia were also identified as key prognostic markers.

**Conclusions:** Machine learning models utilising registry-based administrative data effectively stratify mortality risk in hospitalised SLE patients with high sensitivity. The dominance of pneumonia and renal failure as predictors underscores the critical need for aggressive infection control and renal monitoring in this population.

## Background

Systemic Lupus Erythematosus (SLE) presents a complex array of challenges in clinical outcomes, particularly concerning mortality risk, influenced heavily by heterogeneity in disease expression and the presence of comorbidities. In low-to-middle-income countries, such as those in Southeast Asia, the aggressive phenotype of SLE has been documented, suggesting a higher incidence of premature mortality compared to Western cohorts^1,2^. Hospitalised SLE patients represent a particularly vulnerable demographic, often admitted due to exacerbations of disease manifestations like severe flares, infections, and organ damage, which collectively heighten the risk of in-hospital mortality ^3,4^.

To enhance clinical decision-making and resource allocation, it is essential to accurately identify patients at increased risk of mortality upon admission. Traditional clinical scoring systems, including the SLE Disease Activity Index (SLEDAI) and generalized severity indices like APACHE, are crucial but may require considerable manpower and may not fully encompass the multifaceted nature of patient comorbidities^5,6^. Comorbid conditions, especially infections and renal or cardiovascular disorders, have been identified as significant determinants of survival outcomes in SLE patients. Studies indicate that infections are a predominant cause of morbidity and mortality, particularly among patients receiving immunosuppressive therapy, who face increased risk profiles due to their compromised immune systems^7,8^.

In resource-limited settings, detailed clinical data such as specific immunological markers might not be readily accessible for effective risk stratification. However, administrative data derived from routinely collected sources, such as International Classification of Diseases (ICD-10) codes, offer a practical alternative for risk assessment ^9,10^. The integration of these data sources enables the development of machine learning (ML) approaches which can manage the complex interactions of various risk factors. ML algorithms, such as Random Forest and Extreme Gradient Boosting (XGBoost), can process large, high-dimensional datasets, revealing predictive patterns that traditional statistics may overlook ^5,6^.

The establishment of systems such as the Indonesian Systemic Lupus Erythematosus Regional (ISLET) registry provides a pivotal framework for addressing these challenges. By consolidating clinical and demographic data through platforms like REDCap, ISLET not only standardizes data collection but also supports ongoing research efforts across Southeast Asia^11^. Analyzing data within the ISLET registry represents a crucial opportunity to explore mortality predictors within this locale effectively.

Our proposed study aims to utilize machine learning models to predict in-hospital mortality among SLE patients, examining the efficacy of variables typically available, such as demographics and prevalent ICD-10 coded comorbidities. By leveraging claims-based registry data, our study seeks to establish a scalable, data-driven approach to risk stratification, thereby enhancing the precision of patient management in tertiary care centers.

## Methods

### Study Design and Data Source

This retrospective prognostic study was conducted at Dr. Sardjito General Hospital, a tertiary academic referral center in Yogyakarta, Indonesia. Data were retrieved from the ISLET registry, a standardized web-based platform designed to capture longitudinal clinical data of SLE patients in Indonesia^11^. The study covered hospital admissions from January 1, 2019, to June 30, 2025. The authors accessed the data for research purposes from July 1, 2025 to September 30, 2025. During and after data collection, the authors did not have access to information that could identify individual participants, as all data were anonymized/de-identified prior to analysis. Ethical approval was granted by the Medical and Health Research Ethics Committee of the Faculty of Medicine, Public Health, and Nursing, Universitas Gadjah Mada (Ref: KE-FK-1629-EC-2025), in accordance with the Declaration of Helsinki.

### Cohort Definition and Variable Selection

The study population comprised all patients admitted to the hospital during the specified period with a primary or secondary diagnosis of Systemic Lupus Erythematosus (ICD-10 code M32) or Cutaneous Lupus (ICD-10 code L93). Patients with incomplete administrative records regarding discharge status or admission dates were excluded from the analysis. The primary outcome of interest was in-hospital mortality, classified as a binary variable (survived vs. deceased).

Predictor variables were extracted from the electronic medical records (EMR) integrated into the registry. Demographic features included age and sex. Hospitalization characteristics consisted of length of stay (LOS) and ward type, with the latter categorized into intensive care units and non-intensive general wards. To capture the clinical complexity of the patients, we analyzed the frequency of all secondary ICD-10 diagnosis codes recorded upon admission. Based on this analysis, the ten most frequent comorbidities were selected as binary predictor variables. These included glomerular disorders in systemic connective tissue disorders, unspecified anaemia, urinary tract infections, disorders of plasma-protein metabolism (specifically hypoalbuminemia), hypokalemia, hypo-osmolality/hyponatremia, essential hypertension, pneumonia, unspecified acute kidney failure, and arthritis.

### Data Preprocessing and Machine Learning Setup

Data cleaning and statistical modelling were performed using the R statistical programming environment^12^. Following the machine learning workflow described by Hanif et al. we utilized the tidymodels framework for data partitioning, preprocessing, and model evaluation^13^. Missing values for continuous variables, such as LOS, were imputed using the median value of the cohort. To prepare the data for modelling, continuous variables were normalized to a standard scale. We also addressed multicollinearity by removing predictors with a Spearman’s correlation coefficient greater than 0.7.

A critical challenge in mortality prediction is the class imbalance, where the number of survivors significantly outweighs non-survivors. To mitigate the bias toward the majority class, we applied the Synthetic Minority Over-sampling Technique (SMOTE) to the training data. This technique generates synthetic examples of the minority class (non-survivors) to create a balanced dataset for model training, thereby improving the model’s sensitivity in detecting high-risk patients. The dataset was then randomly split into a training set (75%) and a testing set (25%), stratified by the mortality outcome to ensure a representative distribution of events in both subsets.

### Model Training and Evaluation

We developed and compared three supervised machine learning algorithms to predict in-hospital mortality: Logistic Regression, Random Forest, and Extreme Gradient Boosting (XGBoost). Logistic Regression was employed as a baseline penalized linear model using Elastic Net regularisation (penalty = 0.01, mixing proportion = 0.5), implemented via the glmnet engine. This approach combines L1 (LASSO) and L2 (Ridge) penalties, providing both variable selection and coefficient shrinkage, which is particularly appropriate given the limited sample size relative to the number of predictors. Random Forest, an ensemble learning method robust to overfitting, was implemented using the ranger engine. XGBoost, known for its efficiency in handling large datasets and capturing non-linear relationships, was implemented using the xgboost engine.

To ensure the robustness of the models and prevent overfitting, we performed 10-fold cross-validation on the training set. This process involved partitioning the training data into ten subsets, training the model on nine, and validating it on the remaining one, repeated ten times. The final models were then evaluated on the independent testing set which was not involved in the training process. Model performance was assessed using a comprehensive set of metrics, including the Area Under the Receiver Operating Characteristic Curve (AUC-ROC), sensitivity (recall), specificity, accuracy, and F1-score. Finally, to provide clinical interpretability for the “black-box” tree-based models, we generated Variable Importance Plots (VIP) to identify and rank the clinical features that most significantly contributed to the prediction of mortality.

## Result

### Comorbidity Profile and Baseline Characteristics

The analysis of the study cohort, which included 327 hospitalised patients, revealed a diverse clinical profile defined by the presence of multiple comorbidities. Based on the frequency of ICD-10 codes recorded upon admission, the ten most prevalent comorbidities identified in this population were glomerular disorders in systemic connective tissue disorders, unspecified anaemia, urinary tract infections, disorders of plasma-protein metabolism, hypokalemia, hypo-osmolality and hyponatremia, essential hypertension, pneumonia, acute kidney failure, and arthritis (Figure 1). Glomerular disorders and anaemia were the most dominant conditions, reflecting the systemic nature of the disease in this tertiary referral setting.

**Figure 1.**
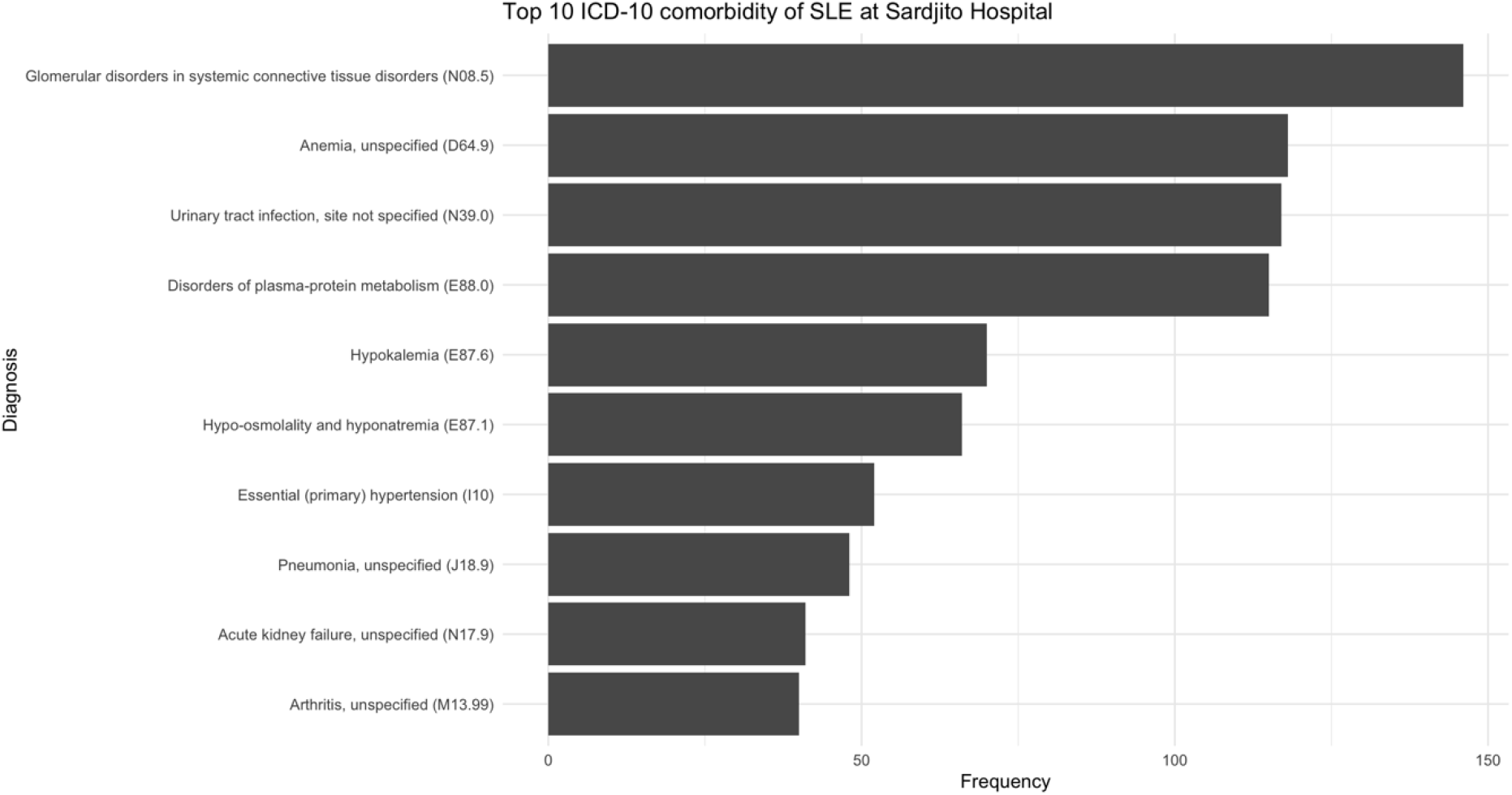
Distribution of the top ten most frequent comorbidities among hospitalised Systemic Lupus Erythematosus (SLE) patients.

The baseline characteristics of the study population, stratified by survival status, are presented in Table 1. The overall in-hospital mortality rate was approximately 7.7% (n = 25). Demographic factors such as age and sex did not show statistically significant differences between survivors and non-survivors; the median age was 30 years for survivors and 34 years for non-survivors (p = 0.13), with a predominantly female population in both groups (>90%). However, significant disparities were observed in hospitalization characteristics. Non-survivors had a significantly longer median length of stay (11.0 days vs. 6.0 days, p < 0.001) and a substantially higher requirement for intensive care services (68% vs. 1.0%, p < 0.001). Regarding specific clinical conditions, non-survivors exhibited a significantly higher prevalence of anaemia (p = 0.008), hypoalbuminemia (p < 0.001), and pneumonia (p < 0.001) compared to survivors. Conversely, comorbidities such as glomerular disorders, urinary tract infections, hypokalemia, hyponatremia, hypertension, and arthritis did not differ significantly between the two groups in the univariate analysis.

**Table 1.** Baseline demographic and clinical characteristics of hospitalised SLE patients stratified by survival status.

| Characteristic | Overall<br>N = 327 | Survivors<br>N = 302 | Non-survivor<br>N = 25 | p-value |
| --- | --- | --- | --- | --- |
| Age, years | 30 (23, 40) | 30 (23, 38) | 34 (25, 47) | 0.13 |
| Sex (% Female) | 26 (8.0%) | 24 (7.9%) | 2 (8.0%) | >0.90 |
| Length of stay, days | 6.0 (3.0, 10.0) | 6.0 (3.0, 10.0) | 11.0 (8.0, 15.0) | <0.001 |
| Ward type (% Intensive) | 20 (6.1%) | 3 (1.0%) | 17 (68%) | <0.001 |
| Glomerular disorder (% Yes) | 111 (34%) | 104 (34%) | 7 (28%) | 0.50 |
| Anaemia (% Yes) | 140 (43%) | 123 (41%) | 17 (68%) | 0.008 |
| Urinary tract infection | 104 (32%) | 92 (30%) | 12 (48%) | 0.070 |
| Hypoalbuminemia | 108 (33%) | 91 (30%) | 17 (68%) | <0.001 |
| Hypokalemia | 50 (15%) | 45 (15%) | 5 (20%) | 0.60 |
| Hyponatremia | 59 (18%) | 52 (17%) | 7 (28%) | 0.20 |
| Hypertension | 77 (24%) | 69 (23%) | 8 (32%) | 0.30 |
| Pneumonia | 60 (18%) | 44 (15%) | 16 (64%) | <0.001 |
| Acute kidney failure | 45 (14%) | 37 (12%) | 8 (32%) | 0.012 |
| Arthritis | 48 (15%) | 44 (15%) | 4 (16%) | 0.80 |

*Data are presented as n (%) for categorical variables and median (IQR) for continuous* *variables. P-values were calculated using the Chi-square test for categorical data and the* *Mann-Whitney U test for continuous data. ICU: Intensive Care Unit; HCU: High Care* *Unit*

### Machine Learning Model Performance

We evaluated the predictive performance of three machine learning algorithms, Logistic Regression, Random Forest, and Extreme Gradient Boosting (XGBoost), using a comprehensive set of metrics. In terms of discrimination capability, all three models demonstrated comparable performance. As illustrated in the Receiver Operating Characteristic (ROC) curves (Figure 2), the XGBoost and Logistic Regression models achieved an identical Area Under the Curve (AUC) of 0.71, marginally outperforming the Random Forest model, which yielded an AUC of 0.70. While the AUC values suggest a moderate level of discrimination consistent with claims-based data, distinct differences emerged when analyzing sensitivity and specificity metrics.

**Figure 2.**
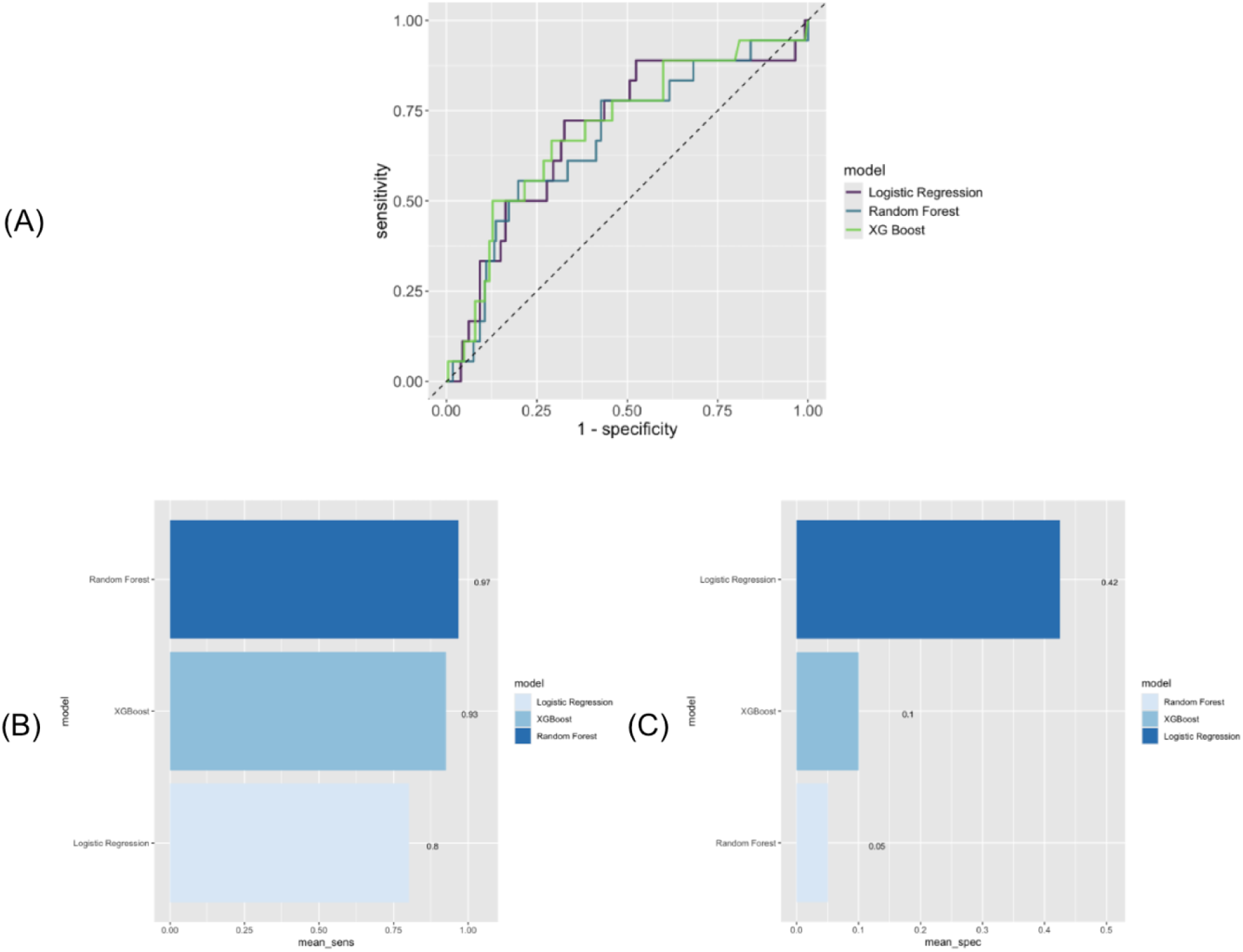
(A) Receiver Operating Characteristic (ROC) curves illustrating the discrimination capability of the three algorithms. (B) Comparison of Mean Sensitivity (Recall) across models. (C) Comparison of Mean Specificity across models

The performance trade-offs between the models were evident in the bar charts illustrating mean sensitivity and specificity. The Logistic Regression model provided the highest specificity (0.42), indicating a better capability to correctly identify survivors, but at the cost of lower sensitivity (0.80). In contrast, the tree-based models prioritized the detection of mortality events. The Random Forest model achieved the highest sensitivity (0.97), followed closely by the XGBoost model (0.93). However, this high sensitivity was accompanied by low specificity for both Random Forest (0.05) and XGBoost (0.10).

### Model Selection and Feature Importance

Despite the trade-off in specificity, the XGBoost model was selected as the optimal algorithm for developing the clinical scoring system. The decision was driven by the clinical priority to minimize false negatives in a mortality prediction context; missing a high-risk patient (Type II error) carries more severe consequences than falsely flagging a low-risk patient. Although Logistic Regression offered a more balanced specificity, its lower sensitivity meant it failed to identify a larger proportion of actual mortality cases compared to XGBoost. Furthermore, XGBoost demonstrated robust performance with an AUC of 0.71, equivalent to Logistic Regression, while offering the capability to model complex, non-linear interactions between variables that linear models might miss.

To interpret the drivers of the XGBoost predictions, we analyzed the feature importance plot (Figure 3), which ranked the variables based on their contribution to the model’s decision-making process. Pneumonia emerged as the single most critical predictor of mortality, exhibiting the highest importance score by a significant margin. This was followed by Acute Kidney Failure and Length of Stay, highlighting the impact of organ failure and prolonged hospitalization on patient outcomes. Other variables included in the top ten features were Age, Hyponatremia, Hypoalbuminemia, Urinary Tract Infection, Glomerular Disorder, Arthritis, and Hypertension. Notably, while variables such as age and hypertension were not statistically significant in the univariate baseline comparison (Table 1), the machine learning algorithm identified them as important contributors in the multivariate context, suggesting their relevant role in the complex interplay of mortality risk factors. These top ten features serve as the foundation for the proposed risk stratification scoring system.

**Figure 3.**
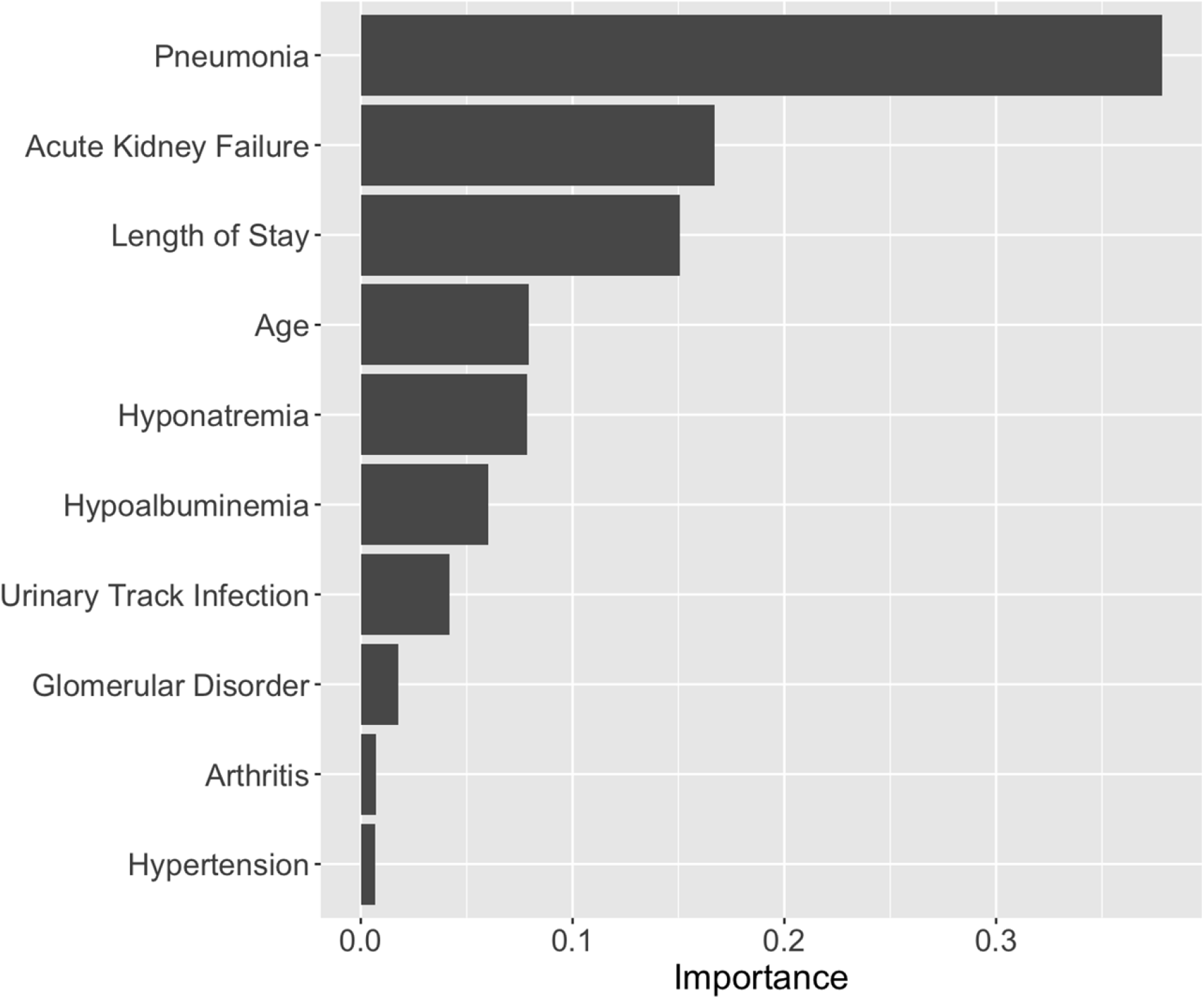
Variable importance plot illustrating the top ten predictors of in-hospital mortality in Systemic Lupus Erythematosus (SLE) patients derived from the Extreme Gradient Boosting (XGBoost) model.

This study was reported in accordance with the Transparent Reporting of a Multivariable Prediction Model for Individual Prognosis or Diagnosis (TRIPOD) guidelines^14^.

## Discussion

This study represents a significant step forward in the application of artificial intelligence for rheumatological care in Southeast Asia, demonstrating the utility of machine learning algorithms in predicting in-hospital mortality among Systemic Lupus Erythematosus patients. By utilising administrative claims data derived from the ISLET registry, our analysis identified that the Extreme Gradient Boosting (XGBoost) model offered the most clinically relevant performance. The model prioritized sensitivity, a crucial attribute for minimizing missed identifications of high-risk patients in acute care settings. The identification of pneumonia, acute kidney failure, and length of stay as the strongest predictors reinforces the established understanding of SLE mortality drivers in Asian populations, while simultaneously highlighting the urgent need for more granular clinical data to refine prognostication.

The emergence of pneumonia as the single most critical predictor of mortality in our XGBoost model (Feature Importance Rank #1) is a finding of profound clinical significance. This aligns with the shifting paradigm of SLE mortality where, unlike in Western cohorts where cardiovascular disease has become the leading cause of death over the last 46 years ^15^, infection remains the predominant cause of mortality in Asian populations. Hamijoyo et al. (2019) reported similar findings in an Indonesian cohort, where infection was a major contributor to morbidity, and Siripaitoon et al. (2014) documented infection as the leading cause of death in Thai SLE patients admitted to intensive care^16,17^.

The pathophysiology behind this high mortality risk is multifactorial. SLE patients face a “double-hit” of immunosuppression: the intrinsic immune dysregulation associated with active lupus and the iatrogenic effects of high-dose corticosteroids or potent immunosuppressants used to control disease flares^18^. In the hospital setting, this susceptibility is often compounded by diagnostic challenges; distinguishing between a pulmonary lupus flare (pneumonitis) and bacterial or opportunistic pneumonia can be difficult, potentially delaying appropriate antibiotic or immunosuppressive therapy. Our findings corroborate observations by Hsu et al. (2005), who noted that severe infections significantly increase the odds of ICU admission and mortality^19^. Consequently, the high feature importance of pneumonia in our model serves as a stark warning to clinicians to maintain a remarkably low threshold for suspecting, diagnosing, and aggressively treating respiratory infections in hospitalised lupus patients to improve survival outcomes.

Acute kidney failure and glomerular disorders were identified as critical predictors, reflecting the aggressive phenotype of Lupus Nephritis (LN) often observed in non-Caucasian populations. Renal involvement is a well-documented independent risk factor for adverse outcomes and mortality^19,20^. In our cohort, the presence of acute kidney injury likely represents either a severe de novo lupus nephritis flare or an acute-on-chronic injury in patients with established renal damage.

Furthermore, our machine learning model identified metabolic derangements, specifically hypoalbuminemia and hyponatremia, as top-ranked features. While often overlooked in traditional scoring systems, these variables act as potent “red flags” in a data-driven model. Hypoalbuminemia in SLE is not merely a marker of nephrotic-range proteinuria but also functions as a negative acute-phase reactant, signaling a state of profound systemic inflammation and catabolism^21^. Similarly, hyponatremia may reflect tubulointerstitial dysfunction, syndrome of inappropriate antidiuretic hormone secretion (SIADH) related to central nervous system involvement, or overall hemodynamic instability. The ability of XGBoost to weigh these metabolic markers heavily suggests that they serve as accessible, low-cost surrogate markers for physiological deterioration that precedes mortality in resource-limited settings.

In developing a predictive model for life-threatening conditions, the balance between sensitivity and specificity is a critical clinical decision. We selected the XGBoost model primarily for its superior sensitivity (0.93) compared to Logistic Regression, despite a trade-off in specificity. In the context of triage in a tertiary referral center, a “false alarm” (false positive), where a stable patient is flagged as high-risk and monitored closely, is clinically acceptable. Conversely, a “missed diagnosis” (false negative), where a dying patient is misclassified as low-risk, can lead to delayed escalation of care and preventable mortality.

This approach aligns with the principles of screening in critical care, where sensitivity is paramount. Wang et al. (2022) previously demonstrated the potential of ensemble methods like Random Forests for classifying severe pneumonia outcomes^22^. Our study extends this by applying gradient boosting methods to a large, contemporary SLE dataset. The model’s ability to capture non-linear interactions between age, comorbidities, and acute diagnoses allows for a more nuanced risk stratification than linear models, making it a valuable tool for early warning systems in emergency departments.

While the XGBoost model performed well, the moderate AUC ~0.71 underscores a critical limitation: the reliance on administrative (ICD-10) claims data. While useful for capturing broad epidemiological trends as noted by Siripaitoon et al. (2014), claims data lack the clinical granularity required for precision rheumatology^17^. Important prognostic variables such as specific autoantibody profiles (e.g., anti-dsDNA, anti-Smith), complement levels (C3/C4), and validated disease activity scores (e.g., SLEDAI-2K) are absent in ICD-10 coding.

This limitation validates the strategic rationale behind the establishment of the ISLET registry^11^. To move beyond moderate predictive accuracy (AUC > 0.80), transitioning from fragmented Electronic Medical Records to a structured, research-grade database is essential. A complete registry that captures longitudinal disease activity, cumulative damage (SLICC/ACR Damage Index), and biopsy-proven nephritis classes would allow machine learning models to distinguish between mortality driven by active lupus versus mortality driven by irreversible organ damage or infection. The integration of such granular data into the ISLET registry is the necessary next step to refine these algorithms for personalized patient care.

The ultimate goal of these predictive models is clinical implementation. Moving forward, integrating these algorithms into clinical practice via web-based tools represents a promising frontier. Similar to the workflow described by Hanif et al. (2024) for stroke prediction, a web-based calculator for SLE mortality could allow clinicians to input key variables readily available at admission (e.g., age, presence of pneumonia, renal function, albumin levels) to receive a real-time risk stratification score^13^. This tool would facilitate evidence-based decision-making, enabling earlier admission to intensive care units, prompt initiation of renal replacement therapy, or more aggressive antimicrobial coverage for high-risk patients, thereby bridging the gap between data science and bedside rheumatology.

Several methodological limitations warrant acknowledgment. First, the tree-based models (Random Forest and XGBoost) were trained using default hyperparameters without systematic tuning, which may have limited their discriminative performance. Future studies should incorporate Bayesian optimisation or grid search cross-validation to maximise model performance. Second, model calibration was not formally assessed; although discrimination AUC was reported, the degree to which predicted probabilities reflect actual event rates remains unknown. Calibration plots and the Brier Score should be incorporated in subsequent validation studies

## Conclusion

In conclusion, this study identifies pneumonia and acute kidney failure as the most significant predictors of in-hospital mortality in Indonesian SLE patients, reflecting the distinct disease patterns in Asian populations. Machine learning models like XGBoost can effectively utilize registry-based claims data to stratify risk with high sensitivity, offering a safety net for high-risk patients. However, to achieve high-precision prognostication, future efforts must focus on enriching the ISLET registry with granular immunological and clinical activity variables, paving the way for AI-assisted precision medicine in lupus care.

## Data Availability

The minimal dataset underlying the findings of this study, consisting of de-identified patient-level administrative variables used in the machine learning analyses, as well as The R analysis code is available from the corresponding author upon reasonable request. Requests will be reviewed to ensure compliance with the ethical approval granted by the Medical and Health Research Ethics Committee, Faculty of Medicine, Public Health and Nursing, Universitas Gadjah Mada (Ref: KE-FK-1629-EC-2025).

## Acknowledgments

None applicable

## Abbreviation list

ACR: American College of Rheumatology
AUC: Area Under the Curve
GLM: Generalized Linear Model
ICD-10: International Classification of Diseases, 10th Revision
IQR: Interquartile Range
ISLET: Indonesian Systemic Lupus Erythematosus Regional
LN: Lupus Nephritis
ML: Machine Learning
REDCap: Research Electronic Data Capture
ROC: Receiver Operating Characteristic
SIADH: Syndrome of Inappropriate Antidiuretic Hormone Secretion
SLE: Systemic Lupus Erythematosus
SLEDAI: Systemic Lupus Erythematosus Disease Activity Index
SLICC: Systemic Lupus International Collaborating Clinics
SMOTE: Synthetic Minority Over-sampling Technique
VIP: Variable Importance Plots
XGBoost: Extreme Gradient Boosting

